# Factors Affecting the Incidence of Stroke at a Young Age: A Philosophical Perspective

**DOI:** 10.1101/2022.07.14.22277618

**Authors:** Titik Agustiyaningsih, Anis Ika Nur Rohmah, Lailatul Azizah

## Abstract

Recently, stroke is a new trend among the young age group in the range of 15-45 years. One of the main complications of this disease is s physical disability, but it also causes communication disorders, emotional disturbances, pain, sleep disturbances, depression, and dysphagia. All of these signs have a major impact on the productivity of the young age group. This article aims to review literatures related to the factors that influence the incidence of stroke at a young age. This study uses a literature study design from 6 databases, namely: Science Direct, ProQuest, Wiley, Sage Pub and Pubmed research. The search used various keyword combinations with the help of Boolean operators, including: “Young Stroke” OR “Young Adults” AND “Risk Factor” OR “Factor”, combined as MESH terms and keywords, and assessment of article quality using the JBI Cohort Studies Cross-Sectional Studies Cohort studies and Case-Control Studies. There were 19 selected articles were analyzed by adjusting the inclusion criteria, namely articles published in the last year, full text accessible, young stroke patient respondents, and discuss the factors that influence the incidence of stroke at a young age. Based on the results of the review, there are several factors that influence the occurance of stroke at a young age including physical factors with a percentage of 85%, lifestyle factors with a percentage of 55%, psychological factors with a percentage of 15%, sex factors with a percentage of 10% and age factors. as much as 5%. Implications in clinical practice include providing psychoeducation as a preventive measure to reduce the incidence of stroke at a young age.

## 1. INTRODUCTION

Stroke according to WHO is defined as clinical signs that occur in global brain dysfunction for more than 24 hours and cause death (Coupland et al., 2017). Today’s stroke, not only occurs in old age but has begun to occur at a young age and has become a new trend among young people (Tsakpounidou et al., 2021). Young strokes are often found in people aged between 15-45 years or under 50 years (Rashid et al., 2020). The wrong lifestyle triggers the risk of stroke at a young age which results in a lack of blood supply entering the brain and causing blockage or rupture of blood vessels (Marbun et al., 2018). In addition, stroke at a young age can cause huge losses such as physical disability at a productive age (Syifa et al., 2017).

Riskesdas 2018 explained that the prevalence of stroke with age > 15 years continued to increase to 10.9 per thousand population. This is also explained by several studies that the percentage of a stroke at the age of 15 years to <24 years is 0.3% and the age of 25 years to <34 years is 0.4%, with a continuous increase at the age of 45 years and over (Putri & Muti, 2010). 2017). National hospitalization data in the United States (NIS) reports an increase in hospitalizations for acute ischemic stroke patients at the age of 18-44 years in 2012 and in 2017 the figure still shows an increase in stroke at a young age (Yahya et al., 2020). In developed countries, the prevalence of stroke at a young age is around 5-10% at the age of <45 years, while in developing countries it is around 19-30% (Hathidara et al., 2019).

Some of the risk factors for stroke at a young age are arteriopathy, heart disorders, chronic systemic conditions, acute systemic disorders, chronic head and neck disorders, acute head and neck disorders, diabetes mellitus, hypertension, smoking, and alcohol consumption (van Alebeek et al., 2018). According to Sinaga & Sembiring (2019), stroke has two risk factors, which are modifiable and non-modifiable. Factors that can be changed include high blood pressure, diabetes mellitus, and dyslipidemia. Age, gender, genetics or race, and acute vascular disorders cannot be changed. Setiawan’s research (2018) also explains that stroke at a young age is caused by several factors that are not good for lifestyle changes, including consuming fast food, smoking, drinking alcohol, rarely exercising, and lack of activity.

Alchuriyah & Wahjuni (2016) stated that women rarely have strokes compared to men who are prone to strokes. However, women in early adulthood between 18-40 years have the same chance of having a stroke as men. This explanation was also presented by the research of Susilawati & Nurhayati (2018) explaining that men are very susceptible to stroke due to their smoking habit which triggers arteriosclerosis and their role as the head of the family which increases stress hormones. In addition, productive age is very busy with all activities so it is rare to spend time exercising lack of rest or sleep, and many thoughts or severe stress which eventually trigger stroke at a young age (Budi et al., 2020).

The increase in stroke at a young age will have a detrimental impact on the physical, psychological, social, and economic (Yaslina et al., 2019). The most frequent impact is the impact on physical, including disability, stress, and depression in people who are dependent on others (Oktari et al., 2020). The psychological impact patients will experience is chaos nges in their emotions, thoughts, and behavior in daily life. Meanwhile, the social impact of the patient will experience negative stigmatization because the community considers a stroke at a young age to be a punishment for the sins committed. The economic impact on patients will experience limitations in terms of expensive care and treatment costs (Yaslina et al., 2019).

The problem of the issue of a stroke at a young age needs to be done more in-depth proof of the factors in the incidence of stroke at a young age which has increased over the past few years. This is because many people, especially young people, always ignore their health and lack public knowledge about the new issues of a stroke at a young age. Therefore, this literature aims to study the philosophy of the factors that influence the incidence of stroke at a young age from a philosophical perspective.

## 2. METHOD

The research method used is an integrative literature review. This review proposes factors that influence stroke at a young age. We systematically searched Science Direct, Pubmed, Proquest, Wiley, Sagepub, and Neliti. The search used various keyword combinations with the help of Boolean operators, including: “Young Stroke” OR “Young Adults” AND “Risk Factor” OR “Factor”, combined as MESH terms and keywords. The inclusion criteria applied in this study were peer-reviewed articles in English and Indonesian that discussed the factors that influence stroke at a young age. Articles published in the last six years (2016-2021). Research studies are conducted in various fields that specifically examine the factors that influence Stroke at a Young Age. This research is quantitative research with cross-sectional studies, cohort studies and case-control studies, and full-text methods. In this study to assess the quality of the journal, researchers used the JBI (Joanna Briggs Institute) assessment instrument. The first author conducted an initial database search and articles for review. We used the PRISMA Flowchart 2009 (Moher et al., 2010) to record the article review and inclusion process (see table 1). An initial search of the four databases yielded 362,039 results. After that, we collect all articles and remove duplicate articles. Sources were excluded by title and abstract if they were not peer-reviewed research studies or related to factors that influence stroke at a young age. The next step is to narrow the selection of articles by year of publication and research context (Figure 1).

**Table 1.** Inclusion and exclusion criteria based on PICOS

| Criteria | Inclusion | Exclusion |
| --- | --- | --- |
| **Population** | Stroke at a young age or adolescents aged 15-45 years or <55 years. | Stroke in the elderly |
| **Intervention** | Stroke factors at a young age | Not including the factors of stroke at a young age |
| **Comparator** | - | - |
| **Outcomes** | Factors that influence stroke at a young age | There is no discussion about the factors that influence stroke at a young age |
| **Study Design** | *Cross-sectional studies, Cohort studies dan Case-Control Studies* | *Randomized Control Trial* |
| **Publication Years** | 2016-2021 | Pre 2016 |
| **Language** | English | Nonenglish language |

**Figure 1.**
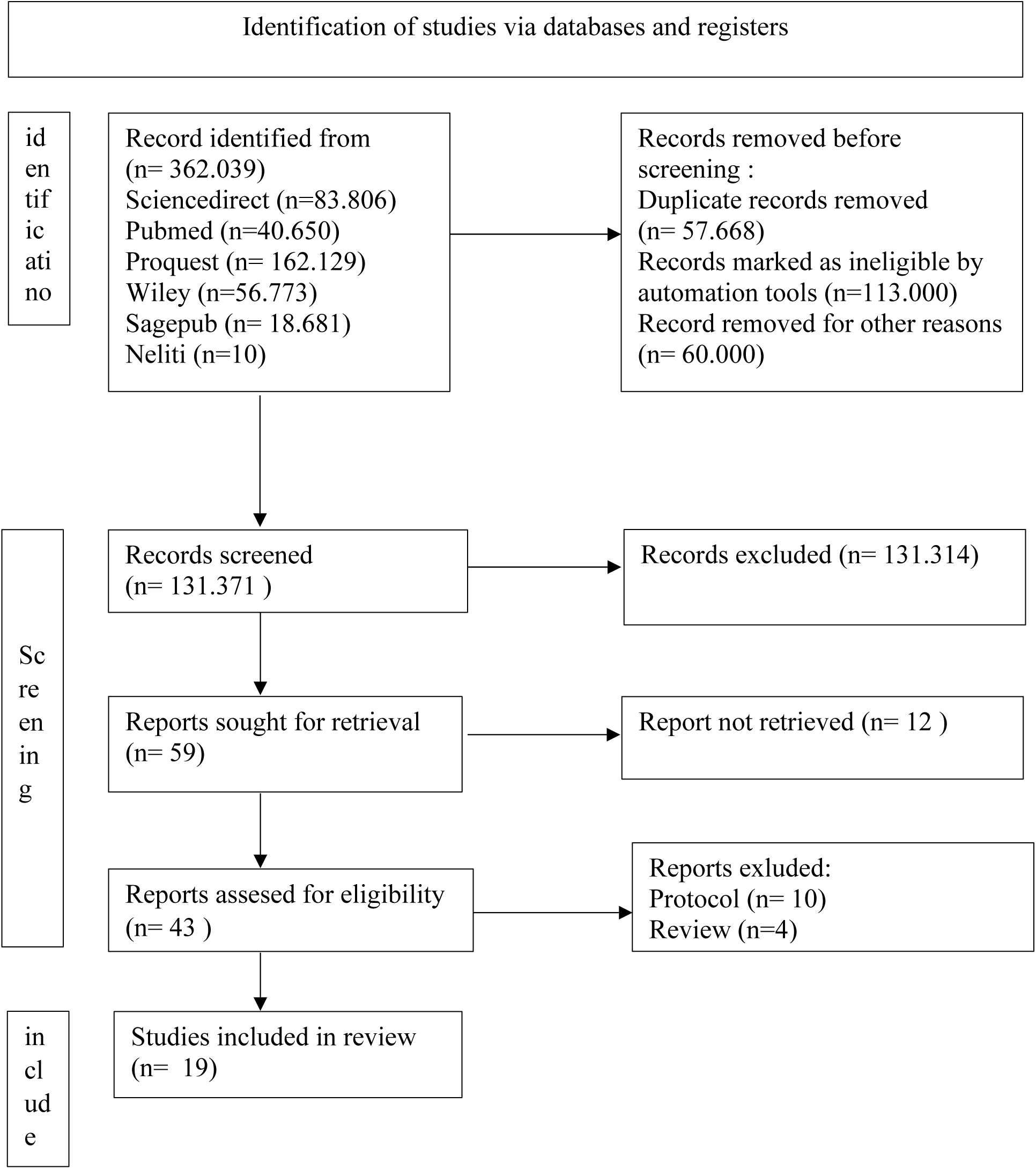
PRISMA Flowchart of Literature Search and Screening Process

## 3. RESULTS AND DISCUSSION

### a. Philosophy of the factors that influence stroke at a young age from an ontology perspective

Stroke at a young age is one of the causes of morbidity and mortality that has an impact on disability and reduced physical productivity at a young age (Mehndiratta & Mehndiratta, 2021). The age most commonly found in young strokes is the age group of 15-45 years (Rashid et al., 2020). WHO clarifies the age standards into 5, the first is young age aged 25-44 years, middle age is 44-60 years old, elderly age is 60-75 years old, senile age is 75-90 years old and long-livers are 90 years and over (Dyussenbayev, 2017). According to the Ministry of Health of the Republic of Indonesia on its official website, depkes.go.id, human ages are divided into several groups, namely toddlers aged 0-5 years, childhood ages 5-11 years, early adolescents 12-16 years old, and late adolescence. 17-25 years old, early adulthood 26-35 years old, late adulthood 36-45 years old, early old age 46-55 years old, late old age 56-65 years old and seniors > 65 years and over (Melanie, 2020).

The increasing prevalence certainly makes young people and the public have to understand the clinical manifestations experienced in stroke at a young age. The clinical manifestations of a stroke at a young age must be watched out for our physical, psychological, and behavioral (Anggriani et al., 2018). Clinical manifestations that often occur in stroke at a young age are feeling tired, feeling sick, neuropathic pain, spasticity, headaches, swollen hands, emotional changes, eye fatigue, slow reading, difficulty recognizing shapes, and aphasia (Stein, 2011). The clinical manifestations are behavioral or social, namely, the patient will be hampered in socializing with others (Millah et al., 2020).

Some of the risk factors for stroke at a young age are arteriopathy, heart disorders, chronic systemic conditions, acute systemic disorders, chronic head and neck disorders, acute head and neck disorders, diabetes mellitus, hypertension, smoking, and alcohol consumption (van Alebeek et al., 2018). According to Sinaga & Sembiring (2019), stroke has two risk factors, which are modifiable and non-modifiable. Factors that can be changed include high blood pressure, diabetes mellitus, and dyslipidemia. The ones that cannot be changed are age, gender, genetics or race, and acute vascular disorders. Setiawan’s research (2018) also explains that stroke at a young age is caused by several factors that are not good for lifestyle changes, including consuming fast food, smoking, drinking alcohol, rarely exercising, and lack of activity.

### b. A philosophy of factors influencing stroke at a young age from an Epistemological perspective

The increasing incidence of stroke at a young age certainly has an adverse impact, one of which causes early disability and sudden death (Susilawati & Nurhayati, 2018). In addition to disability and paralysis, stroke can cause communication disorders, emotional disturbances, pain, sleep disturbances, depression, and dysphagia. Even the level of dependence on others increases loses confidence and loses enthusiasm for life (Karunia., 2016). According to the Barthel index assessment, patients with stroke who have a high level of dependence have a significant relationship to emotional distress (Oktari et al., 2020).

The impact on the quality of life of young stroke patients is lower than that of old age strokes because they are still productive at work, educated, and caring for their families, so there is still much to be accomplished in their lives. Research by Bartholomé & Winter (2020) explains that the most influential quality of life at a young age is physical, emotional, and social. In terms of work, young stroke patients will lose their jobs and even often take time off work due to poor body function (Akinwuntan et al., 2021). The impacts that occur on family members include financial problems, loss of roles or shifts in family roles, and even damage to family relationships. Anxiety and worry also occur in the family and caregivers of stroke patients experiencing emotional stress during caring for stroke patients (E. Y. H. Tang et al., 2020).

To avoid this, the need for prevention efforts to reduce the incidence of stroke at a young age. Based on research (Budi et al., 2020) explains that these promotive and preventive efforts can increase public awareness to control hypertension, exercise regularly, and reduce consumption of fatty foods by conducting health education and discharge planning for stroke patients and their families and communities of productive age. In addition, research (Yahya et al., 2020) explains that the prevention of stroke at a young age is divided into two primordial prevention and primary prevention. Primordial prevention includes smoking cessation, weight management, regular physical activity, and a healthy diet whereas primary prevention consists of statin therapy and therapy to lower lipoproteins.

### c. A philosophy of factors influencing stroke at a young age from an axiological perspective

Based on a literature review of several studies (Table 2), this discussion explains the factors that influence stroke at a young age through 20 journals that have been studied where there are physical factors, psychological factors, lifestyle factors, age factors, and gender factors.. The discussion regarding the results of this literature study is as follows:

**Table 2.**
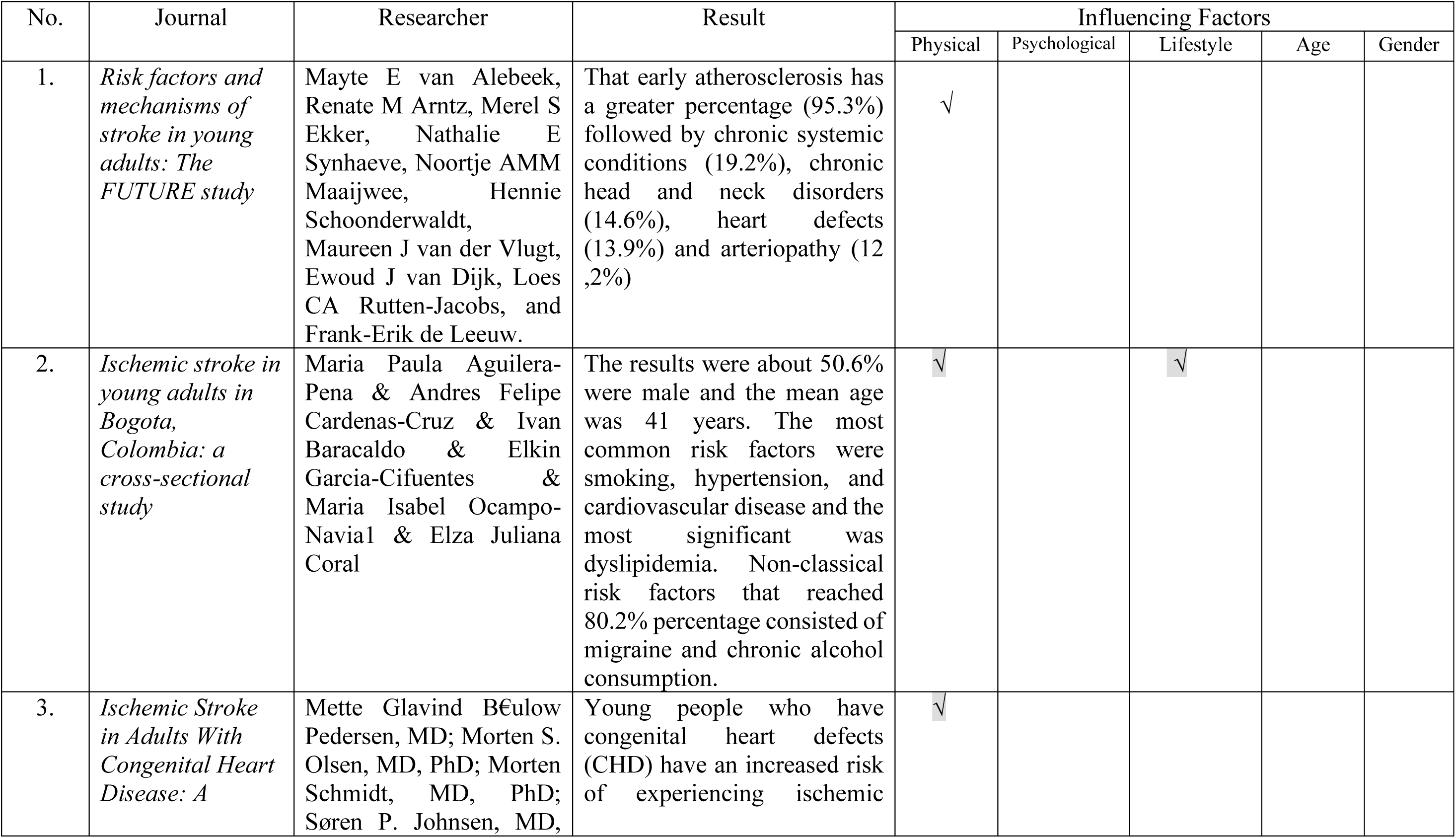
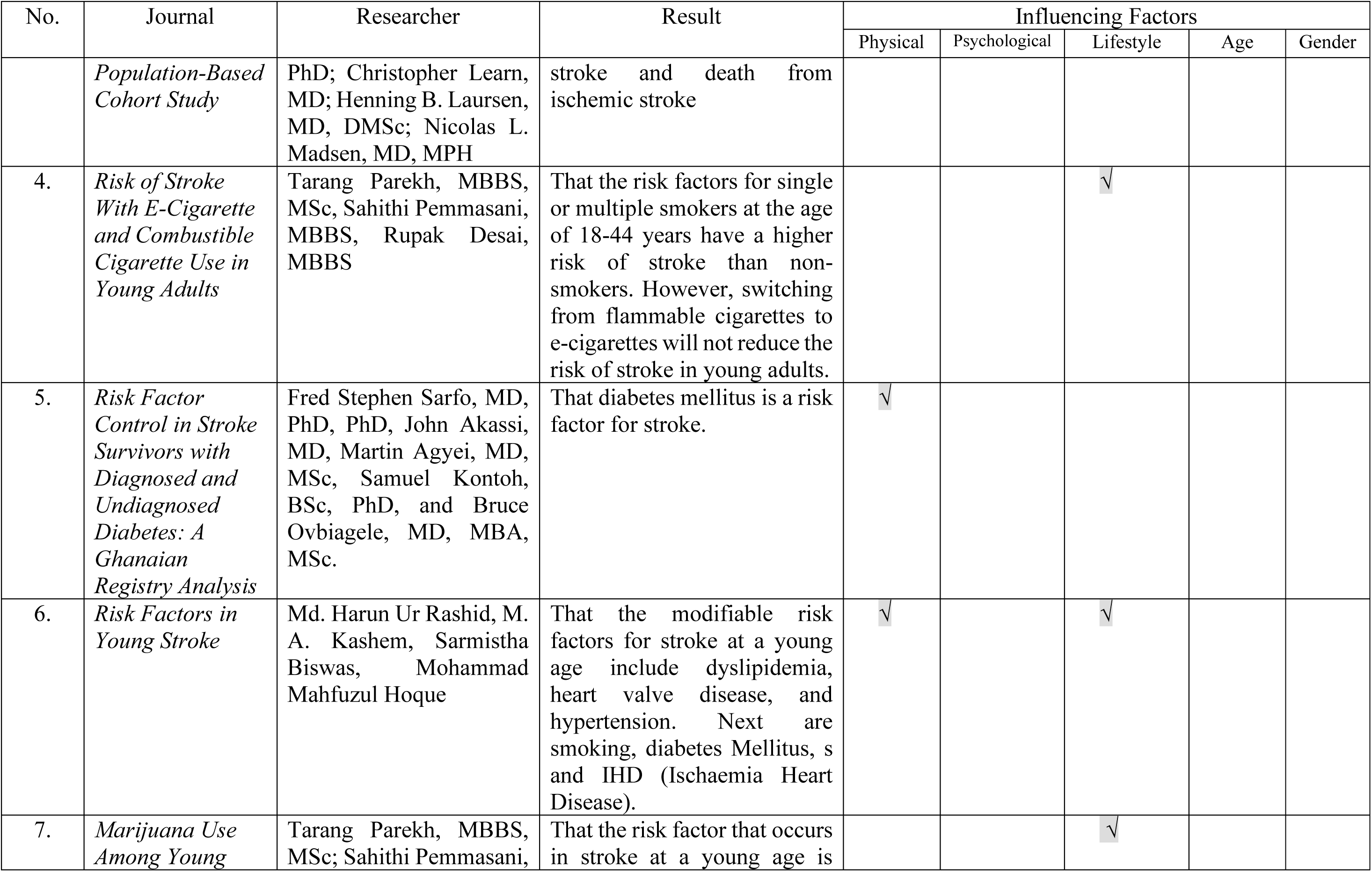
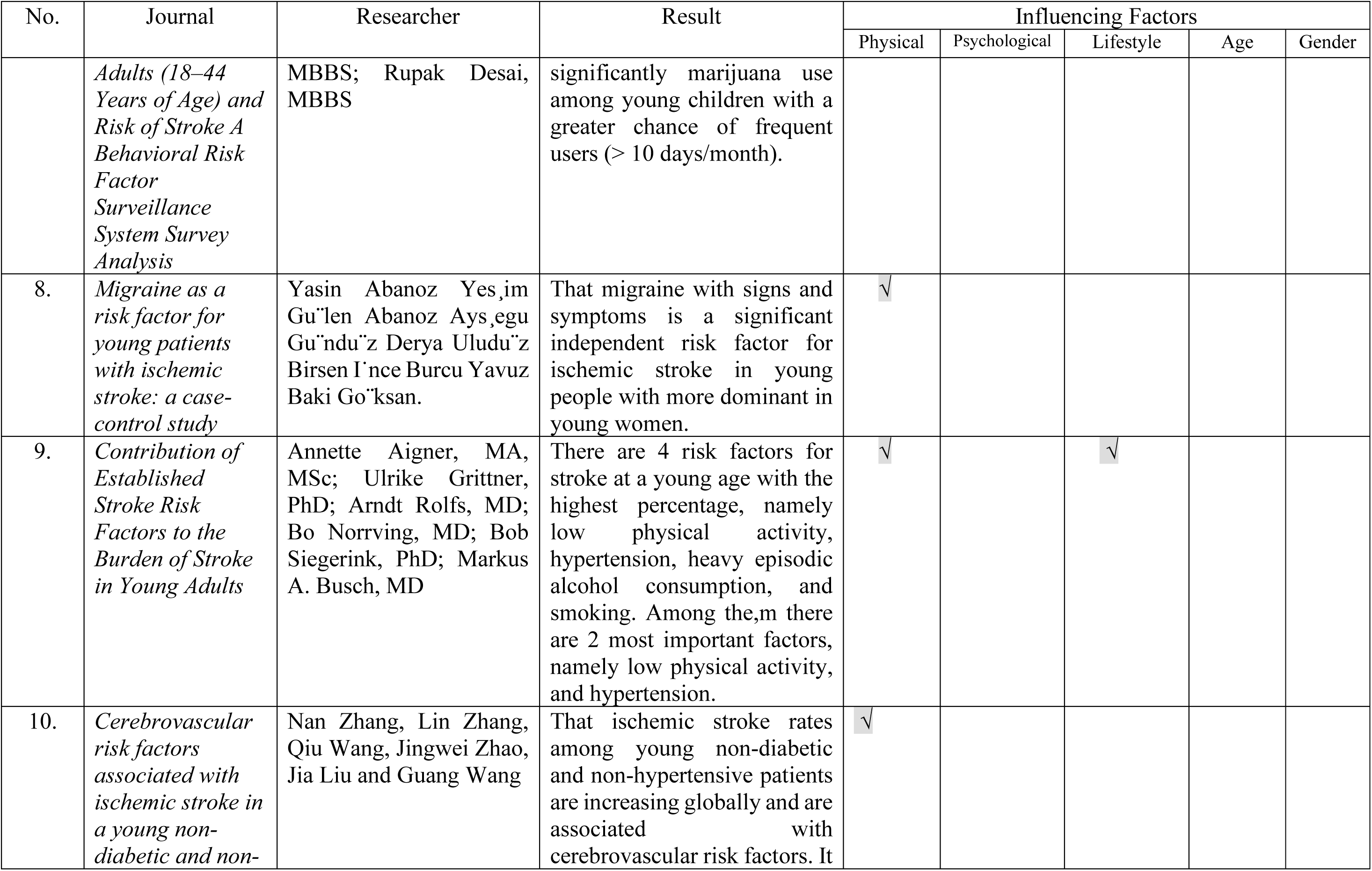
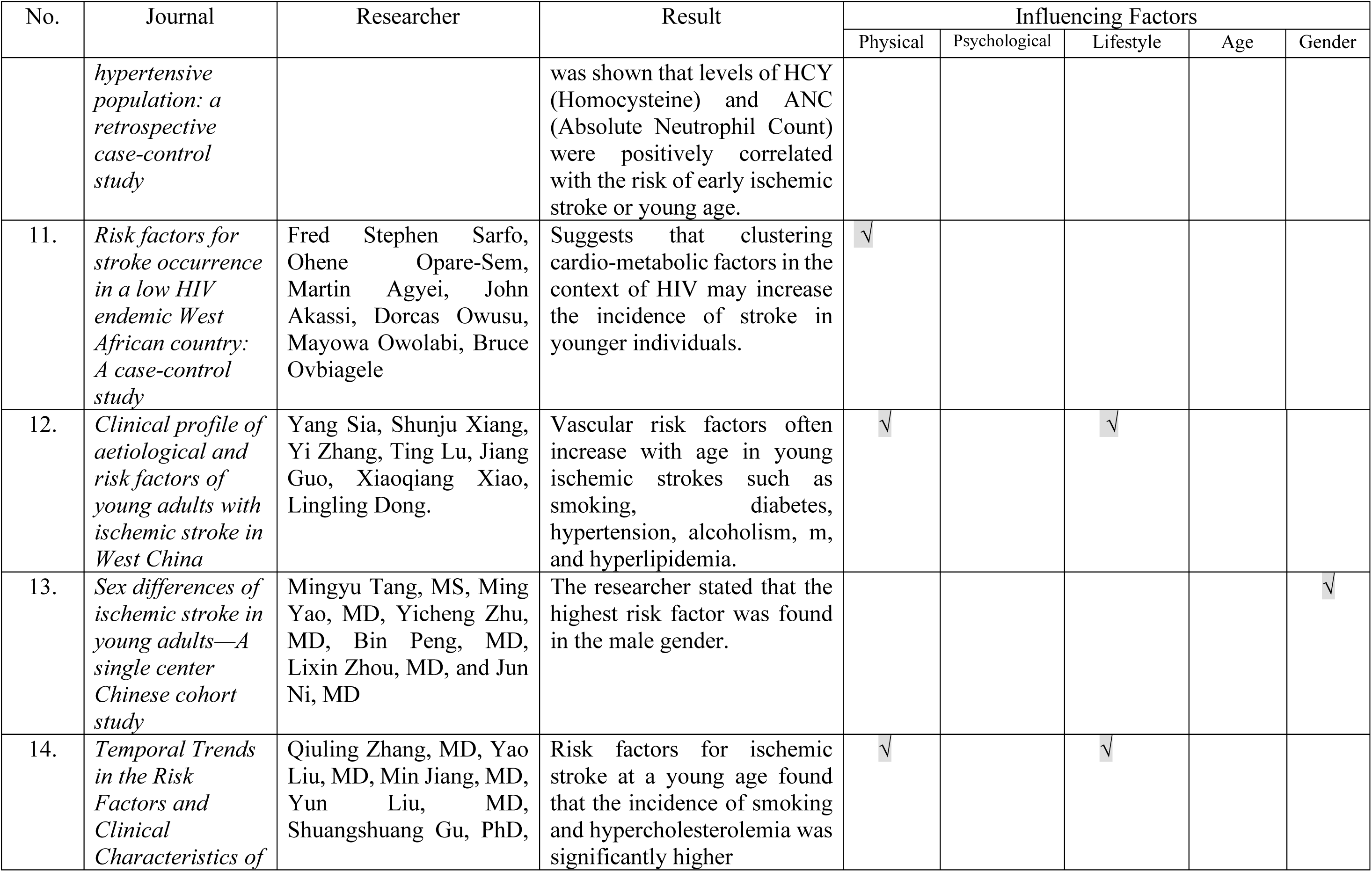
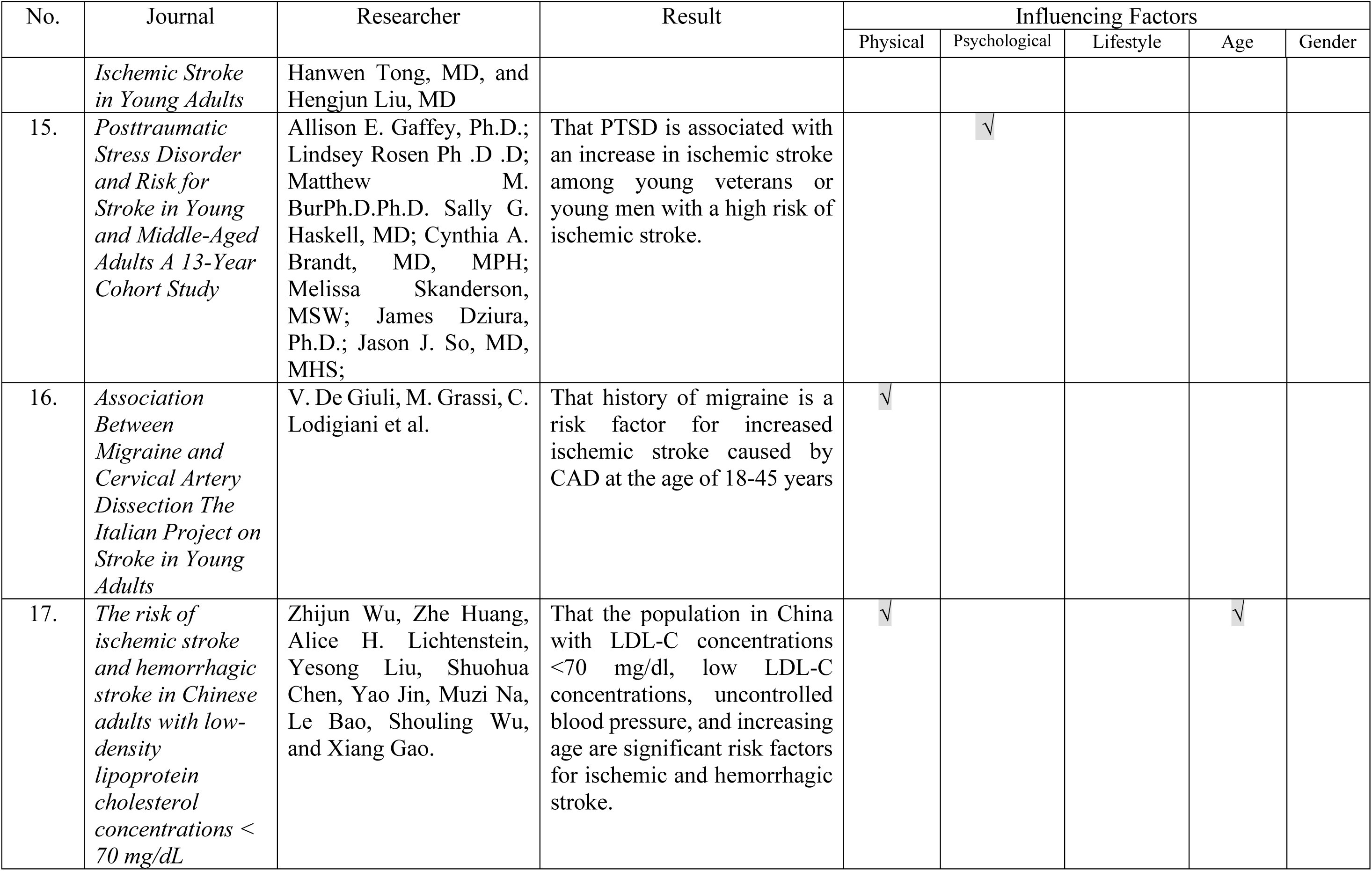
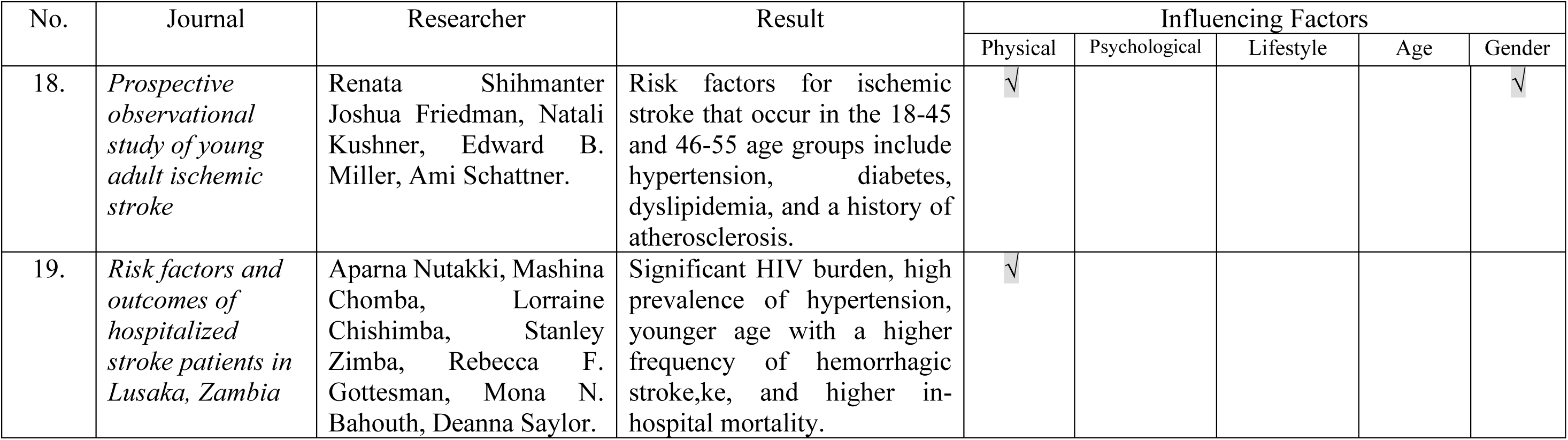
The results of the review of articles based on the Axiological perspective approach (n=19)

| No. | Journal | Researcher | Result | Influencing Factors |  |  |  |  |
| --- | --- | --- | --- | --- | --- | --- | --- | --- |
|  |  |  |  | Physical | Psychological | Lifestyle | Age | Gender |
| 1. | *Risk factors and mechanisms of stroke in young adults: The FUTURE study* | Mayte E van Alebeek, Renate M Arntz, Merel S Ekker, Nathalie E Synhaeve, Noortje AMM Maaijwee, Hennie Schoonderwaldt, Maureen J van der Vlugt, Ewoud J van Dijk, Loes CA Rutten-Jacobs, and Frank-Erik de Leeuw. | That early atherosclerosis has a greater percentage (95.3%) followed by chronic systemic conditions (19.2%), chronic head and neck disorders (14.6%), heart defects (13.9%) and arteriopathy (12,2%) | √ |  |  |  |  |
| 2. | *Ischemic stroke in young adults in Bogota, Colombia: a cross-sectional study* | Maria Paula Aguilera-Pena & Andres Felipe Cardenas-Cruz & Ivan Baracaldo & Elkin Garcia-Cifuentes & Maria Isabel Ocampo-Navia1 & Elza Juliana Coral | The results were about 50.6% were male and the mean age was 41 years. The most common risk factors were smoking, hypertension, and cardiovascular disease and the most significant was dyslipidemia. Non-classical risk factors that reached 80.2% percentage consisted of migraine and chronic alcohol consumption. | √ |  | √ |  |  |
| 3. | *Ischemic Stroke in Adults With Congenital Heart Disease: A* | Mette Glavind Bøulow Pedersen, MD; Morten S. Olsen, MD, PhD; Morten Schmidt, MD, PhD; Søren P. Johnsen, MD, | Young people who have congenital heart defects (CHD) have an increased risk of experiencing ischemic | √ |  |  |  |  |
|  |  |  |  | Physical | Psychological | Lifestyle | Age | Gender |
|  | *Population-Based Cohort Study* | PhD; Christopher Learn, MD; Henning B. Laursen, MD, DMSc; Nicolas L. Madsen, MD, MPH | stroke and death from ischemic stroke |  |  |  |  |  |
| 4. | *Risk of Stroke With E-Cigarette and Combustible Cigarette Use in Young Adults* | Tarang Parekh, MBBS, MSc, Sahithi Pemmasani, MBBS, Rupak Desai, MBBS | That the risk factors for single or multiple smokers at the age of 18-44 years have a higher risk of stroke than non-smokers. However, switching from flammable cigarettes to e-cigarettes will not reduce the risk of stroke in young adults. |  |  | √ |  |  |
| 5. | *Risk Factor Control in Stroke Survivors with Diagnosed and Undiagnosed Diabetes: A Ghanaian Registry Analysis* | Fred Stephen Sarfo, MD, PhD, PhD, John Akassi, MD, Martin Agyei, MD, MSc, Samuel Kontoh, BSc, PhD, and Bruce Ovbiagele, MD, MBA, MSc. | That diabetes mellitus is a risk factor for stroke. | √ |  |  |  |  |
| 6. | *Risk Factors in Young Stroke* | Md. Harun Ur Rashid, M. A. Kashem, Sarmistha Biswas, Mohammad Mahfuzul Hoque | That the modifiable risk factors for stroke at a young age include dyslipidemia, heart valve disease, and hypertension. Next are smoking, diabetes Mellitus, s and IHD (Ischaemia Heart Disease). | √ |  | √ |  |  |
| 7. | *Marijuana Use Among Young* | Tarang Parekh, MBBS, MSc; Sahithi Pemmasani, | That the risk factor that occurs in stroke at a young age is |  |  | √ |  |  |
|  |  |  |  | Physical | Psychological | Lifestyle | Age | Gender |
|  | *Adults (18–44 Years of Age) and Risk of Stroke A Behavioral Risk Factor Surveillance System Survey Analysis* | MBBS; Rupak Desai, MBBS | significantly marijuana use among young children with a greater chance of frequent users (> 10 days/month). |  |  |  |  |  |
| 8. | *Migraine as a risk factor for young patients with ischemic stroke: a case-control study* | Yasin Abanoz Yesim Gu'len Abanoz Ayselgu Gu'ndu'z Derya Uludu'z Birsen I'nce Burcu Yavuz Baki Go'ksan. | That migraine with signs and symptoms is a significant independent risk factor for ischemic stroke in young people with more dominant in young women. | √ |  |  |  |  |
| 9. | *Contribution of Established Stroke Risk Factors to the Burden of Stroke in Young Adults* | Annette Aigner, MA, MSc; Ulrike Grittner, PhD; Arndt Rolfs, MD; Bo Norrving, MD; Bob Siegerink, PhD; Markus A. Busch, MD | There are 4 risk factors for stroke at a young age with the highest percentage, namely low physical activity, hypertension, heavy episodic alcohol consumption, and smoking. Among them there are 2 most important factors, namely low physical activity, and hypertension. | √ |  | √ |  |  |
| 10. | *Cerebrovascular risk factors associated with ischemic stroke in a young non-diabetic and non-* | Nan Zhang, Lin Zhang, Qiu Wang, Jingwei Zhao, Jia Liu and Guang Wang | That ischemic stroke rates among young non-diabetic and non-hypertensive patients are increasing globally and are associated with cerebrovascular risk factors. It | √ |  |  |  |  |
|  |  |  |  | Physical | Psychological | Lifestyle | Age | Gender |
|  | *hypertensive population: a retrospective case-control study* |  | was shown that levels of HCY (Homocysteine) and ANC (Absolute Neutrophil Count) were positively correlated with the risk of early ischemic stroke or young age. |  |  |  |  |  |
| 11. | *Risk factors for stroke occurrence in a low HIV endemic West African country: A case-control study* | Fred Stephen Sarfo, Ohene Opere-Sem, Martin Agyei, John Akassi, Dorcas Owusu, Mayowa Owolabi, Bruce Ovbiagele | Suggests that clustering cardio-metabolic factors in the context of HIV may increase the incidence of stroke in younger individuals. | √ |  |  |  |  |
| 12. | *Clinical profile of aetiological and risk factors of young adults with ischemic stroke in West China* | Yang Sia, Shunju Xiang, Yi Zhang, Ting Lu, Jiang Guo, Xiaoqiang Xiao, Lingling Dong. | Vascular risk factors often increase with age in young ischemic strokes such as smoking, diabetes, hypertension, alcoholism, m, and hyperlipidemia. | √ |  | √ |  |  |
| 13. | *Sex differences of ischemic stroke in young adults—A single center Chinese cohort study* | Mingyu Tang, MS, Ming Yao, MD, Yicheng Zhu, MD, Bin Peng, MD, Lixin Zhou, MD, and Jun Ni, MD | The researcher stated that the highest risk factor was found in the male gender. |  |  |  |  | √ |
| 14. | *Temporal Trends in the Risk Factors and Clinical Characteristics of* | Qiuling Zhang, MD, Yao Liu, MD, Min Jiang, MD, Yun Liu, MD, Shuangshuang Gu, PhD, | Risk factors for ischemic stroke at a young age found that the incidence of smoking and hypercholesterolemia was significantly higher | √ |  | √ |  |  |
|  |  |  |  | Physical | Psychological | Lifestyle | Age | Gender |
|  | *Ischemic Stroke in Young Adults* | Hanwen Tong, MD, and Hengjun Liu, MD |  |  |  |  |  |  |
| 15. | *Posttraumatic Stress Disorder and Risk for Stroke in Young and Middle-Aged Adults A 13-Year Cohort Study* | Allison E. Gaffey, Ph.D.; Lindsey Rosen Ph.D.; Matthew M. BurPh.D.Ph.D. Sally G. Haskell, MD; Cynthia A. Brandt, MD, MPH; Melissa Skanderson, MSW; James Dziura, Ph.D.; Jason J. So, MD, MHS; | That PTSD is associated with an increase in ischemic stroke among young veterans or young men with a high risk of ischemic stroke. |  | √ |  |  |  |
| 16. | *Association Between Migraine and Cervical Artery Dissection The Italian Project on Stroke in Young Adults* | V. De Giuli, M. Grassi, C. Lodigiani et al. | That history of migraine is a risk factor for increased ischemic stroke caused by CAD at the age of 18-45 years | √ |  |  |  |  |
| 17. | *The risk of ischemic stroke and hemorrhagic stroke in Chinese adults with low-density lipoprotein cholesterol concentrations < 70 mg/dL* | Zhijun Wu, Zhe Huang, Alice H. Lichtenstein, Yesong Liu, Shuohua Chen, Yao Jin, Muzi Na, Le Bao, Shouling Wu, and Xiang Gao. | That the population in China with LDL-C concentrations <70 mg/dl, low LDL-C concentrations, uncontrolled blood pressure, and increasing age are significant risk factors for ischemic and hemorrhagic stroke. | √ |  |  | √ |  |
|  |  |  |  | Physical | Psychological | Lifestyle | Age | Gender |
| 18. | *Prospective observational study of young adult ischemic stroke* | Renata Shihmanter Joshua Friedman, Natali Kushner, Edward B. Miller, Ami Schattner. | Risk factors for ischemic stroke that occur in the 18-45 and 46-55 age groups include hypertension, diabetes, dyslipidemia, and a history of atherosclerosis. | √ |  |  |  | √ |
| 19. | *Risk factors and outcomes of hospitalized stroke patients in Lusaka, Zambia* | Aparna Nutakki, Mashina Chomba, Lorraine Chishimba, Stanley Zimba, Rebecca F. Gottesman, Mona N. Bahouth, Deanna Saylor. | Significant HIV burden, high prevalence of hypertension, younger age with a higher frequency of hemorrhagic stroke, and higher in-hospital mortality. | √ |  |  |  |  |

#### a. Physical Factor

This study stated that most of the 20 selected journals contained 17 journals that described physical factors. One of the comorbidities that mostly affects stroke at a young age with a percentage of as much as 85%. The journal describes the comorbidities that affect stroke at a young age ranging from premature atherosclerosis, chronic systemic conditions, head and neck disorders, and heart disorders. Additional factors include arteriopathy, genetic disorders, hypertension, migraine, dyslipidemia, diabetes, high cholesterol, heart failure, obesity, HIV with stroke, cardioembolism, m and heart valve disease. Comorbidities significantly increase mortality in post-stroke patients and mortality in the general population.

The effects of these comorbidities are more common in stroke patients under the age of 60 or younger. Patients who have comorbidities generally have more severe strokes that affect patient disability and death (Corraini et al., 2018). Comorbidities or what are known as comorbidities are an important factor in the burden of stroke. According to the Charlson Comorbidity Index (CCI), it is identified that these comorbidities are very common in stroke patients so they require more complex treatment and recovery due to comorbidities. Stroke patients who have a high CCI (Charlson Comorbidity Index) score increase the risk of poor outcomes and even death (Nelson et al., 2017).

Based on this study, the most common physical factors in young stroke patients based on the journal were hypertension, diabetes, dyslipidemia, early atherosclerosis, and heart disease.

##### 1) Hypertension

Hypertension or high blood pressure is a major risk factor for cardiovascular and cerebrovascular diseases, one of which is stroke (Cho & Yang, 2018). As many as 40% of stroke patients suffer from hypertension, this is similar to other studies which state that 74 patients with hypertension suffer from stroke and about 34% of patients do not realize that they have hypertension and 60.7% undergo irregular treatment (Rashid et al., 2020). The prevalence of hypertension based on data from the Indonesian Ministry of Health states that hypertension in Indonesia reaches 31.7% of the population aged 18 years and over and about 60% of people with hypertension end up with stroke (Yonata & Pratama, 2016). The mechanism of the pathophysiological process in hypertension so that it becomes a stroke is very significant. First, chronic hypertension causes major changes in cerebral circulation that have an impact on cerebral blood flow, namely structural changes. Hypertrophy of the vessel wall is defined as an increase in wall thickness, an increase in the cross-sectional area of the war, l, l, or an induced increase in the wall-to-lumen ratio. Hypertrophy induced by hypertension and remodeling (changes) is thought to be both protective and detrimental to cerebral arterioles. Thus, increased intraluminal pressure, as in chronic hypertension, increases wall tension. In the circulation in the brain, that cerebral arterioles in the SHRSP will experience a reduction in radius and an increase in wall thickness, thereby increasing the vasoconstrictive response. Vasoconstrictive stimulation during hypertension will cause narrowing of cerebral arterioles and increase sympathetic nerve density or catecholamine flow and dilation of cerebral arterioles (Cipolla et al., 2018).

##### 2) Diabetes Mellitus

Another comorbid factor that triggers stroke at a young age is diabetes mellitus. Patients with diabetes mellitus have a 1.5-3 fold risk of stroke with a high mortality rate compared to patients without diabetes. The prevalence of diabetes mellitus with the incidence of stroke at a young age according to the research by Echouffo-Tcheugui (2018) is 40% for those aged <50 years. This metabolic disorder is caused by abnormal fat accumulation in the arteries so that patients with hyperglycemia (high blood glucose) have a 1.5-fold increased risk of stroke. Complications that occur in vascular diabetes are divided into two, namely microvascular complications consisting of neuropathy, retinopathy, and diabetic nephropathy while macrovascular complications include stroke, peripheral arterial disease, and coronary blood vessels (Alloubani et al., 2018). The pathophysiological mechanisms that occur in this risk factor are firstly increased oxidative stress resulting from hyperglycemia it willwhichead to pathological processes involved in microvascular complications associated with diabetes. So as a result of the increased reactive oxygen species (ROS) it will inhibit the action of glyceraldehyde 3-phosphate dehydrogenase (GADPH) or the key enzyme of glycolysis. Free radicals induce DNA cleavage and reactive oxygen species (ROS) activate the enzyme poly DNA (ADP-Ribose) Polymerase (PARP). This PARP will modify GADPH and inhibit all its activities. The subsequent mechanism process results in the accumulation of glycolytic intermediates and GADPH to form 5 pathogenic pathways that contribute to endothelial dysfunction and complications in diabetes mellitus. Chronic hyperglycemia-induced vasculopathy associated with endothelial damage results in accelerated atherosclerosis in diabetes mellitus. So that a higher prevalence of cardiovascular disease including stroke often occurs in diabetes mellitus (Tun et al., 2017).

##### 3) Dyslipidemia

Dyslipidemia increases the risk of stroke recurrence in stroke patients who have large artery atherosclerosis. However, this does not happen in all patients. One study stated that the stroke recurrence rate was significantly higher in patients with atherogenic dyslipidemia compared to patients who did not have this condition. The country in Estonia also explained that young patients with the first stroke and recurrent stroke showed a high prevalence in patients with dyslipidemia, the percentages were around 45.5% and 47.9% (Sarecka-Hujar & Kopyta, 2020). This explanation is the same as the study according to Matuja et al (2020) that dyslipidemia is a risk factor for stroke originating from atherosclerosis, which impairs blood pressure regulation that causes hypertension. Other trigger factors that raise the possibility of dyslipidemia include smoking, alcohol consumption, in, diabetes mellitus.

##### 4) Early Atherosclerosis

Early atherosclerosis is the most widely described risk factor for stroke at a young age from the results of this journal study. A research study by van Alebeek et al., (2018) explains that risk factors for early atherosclerosis are more commonly found in stroke patients aged > 35 years compared to patients with younger ages < 35 years according to TOAST criteria with percentages of 96.9% and 89, respectively., 0%. In general, with increasing age in young stroke patients, the burden of vascular risk factors increases. Hypertension, diabetes, and hyperlipidemia are precursors to atherosclerosis. This factor is very important because young stroke patients who have atherosclerosis have a higher mortality percentage than patients who do not have this condition (Stack & Cole, 2018).

##### 5) Heart Disease

Comorbid factors that often occur in stroke at a young age are heart disease. Heart disease is a very common cause of cardioembolism in young stroke patients. Structural abnormalities of the heart are a potential cause of heart disease at a young age. The percentage of ischemic stroke incidence in young people with CHD is 0.7%, while in the general population it is 0.1%. The Baltimore-Washington Young Stroke Study identified bacterial endocarditis as the most common cause of heart disease. Heart defects have a higher relative risk of between 10 and 100 times greater in young stroke patients than those without heart defects (Stack & Cole, 2018).

#### b. Psychological Factors

Based on the research results, the factors that influence stroke at a young age are psychological. The most psychological factors in this study were stress, emotional disturbances, and the, the incidence of PTSDPsychologicalll factors that play an important role in the predisposition to multifactorial stroke (Polivka et al., 2019)

##### 1) Stress

One factor that influences the occurrence stra oke at a young age is stress. Stress is a modifiable risk factor for stroke at a young age. Stress often occurs in someone who has a perfectionist and obsessive personality. This stress is related to psychosocial issues ranging from family problems, work problems ad, and life events that eventually cause stress and even depression. Another study also concluded that these psychological factors increase the risk of stroke by 39%. (Polivka et al., 2019). The pathophysiology of stress or depression in stroke is highly multifactorial, involving a combination of ischemic neurobiological dysfunction and psychosocial stress. Damage to the brainstem pathways of the frontal-basal gachangesges in the monoaminergic neurotransmitter system. According to evidence, there is a relationship between the neuroinflammatory response to acute ischemic stroke, activation of stress in the hypothalamic-pituitary-adrenal (HPA), and decreased adaptive response (neurogenesis) in mitochondrial dysfunction (Villa et al., 2018).

##### 2) Emotional Disorder

According to Zevon and Tellegen, the category of emotions is divided into 5 categories, namely category 1 is nervous, depressed, afraid, category 2 is sad, category 3 is angry and annoyed, category 4 is angry with oneself, guilty and dissatisfied with oneself and category 5 is calm. and satisfied. Several studies have shown that harsh life events and a lot of stress that cause emotional disturbances are risk factors for stroke and are interrelated with psychological factors (Prasad et al., 2020).

##### 3) PTSD

The results of this study explain that Post Traumatic Stress Disorder (PTSD) is a risk factor for stroke at a young age that often occurs. Post-traumatic stress disorder (PTSD) is a treatable mental health condition for young people and begins with exposure to severe psychological stress or trauma. Post Traumatic Stress Disorder (PTSD) is associated with an increased risk of ischemic stroke at a young age and an increased risk of early hemorrhagic stroke. In addition, patients with Post Traumatic Stress Disorder (PTSD) often have many risk factors for stroke, one of which is hypertension (Gaffey et al., 2021).

#### c. Lifestyle Factors (Lifestyle)

The incidence of stroke at a young age is also influenced by lifestyle factors. Unfavorable lifestyle factors can trigger strokes at a young age (Polivka et al., 2019). Some of the results of this research journal that the most common lifestyle factors in stroke at a young age include smoking, alcohol consumption, marijuana consumption, poor diet, low activity, and use of oral contraceptives in women.

##### a) Smoking

Smoking habits are the biggest trigger for stroke at a young age. Although smoking has no significant relationship with the incidence of stroke, it turns out that smoking is a real cause of stroke at a young age (Simbolon et al., 2018). The percentage of stroke incidence at a young age in smokers is 83% higher than n non-smokers (Parekh et al., 2020). A smoker has a seven-fold risk of stroke than a non-smoker. Smoking can also cause rapid cerebral vasoconstriction, an increase in above-average arterial pressure and an increase in blood glucose and is associated with increased blood viscosity and dyslipidemia (Dinh et al., 2019). The pathophysiology of smoking causing a stroke is not fully understood. However, there is a dose relationship between the number of cigarettes smoked and an increased risk of stroke. Smoking produces many pathologies that are closely related to a high risk of stroke. Smoking can impair vasodilator function and decrease the availability of nitrous oxide and major dilators. Smoking can also cause rapid cerebral vasoconstriction, an increase in above-average arterial pressure, e and an increase in blood glucose and is associated with increased blood viscosity and dyslipidemia (Dinh et al., 2019).

##### b) Alcohol consumption

Very high alcohol consumption can be a trigger for stroke at a young age. Research in Sweden Falkstedt et al., (2017) state that there is a significant relationship between high alcohol consumption and the incidence of stroke, whether drinking beer, wine, and other types of liquor. Consuming excessive alcohol will damage the nervous system in the brain and have neurological and psychological side effects (Dguzeh et al., 2018). Excessive alcohol consumption will damage the nervous system in the brain and have neurological and psychological side effects. When affected by alcohol, the hippocampus, which controls memory, can experience memory loss or memory loss. The hypothalamus coordinates important activities of the pituitary gland and autonomic nervous system and controls body temperature, hunger, thirst, st and other homeostatic systems. However, after drinking too much alcohol, hunger, thirst, blood pressure, and the urge to urinate increase while body temperature decreases. The medulla oblongata is also affected by the AUD which is responsible for involuntary functions such as body temperature regulation and breathing. When the medulla is affected, body temperature will decrease and breathing will be depressed (Dguzeh et al., 2018).

##### c) Marijuana consumption

The prevalence of marijuana use was significantly higher in the age of 18-24 years in men, which was about 63.3%. Marijuana use in the United States has increased at a young age, allowing the risk of stroke at young age due to the high use of marijuana. Excessive marijuana consumption (>10 days/month) has a very high risk of stroke at a young age (Parekh et al., 2020). Stroke associated with drug or drug abuse causes direct damage to the blood vessels of the brain either directly or indirectly. It also affects other organs such as the heart and cerebral circulation. The mechanism of action of marijuana begins with its lipophilic nature and cadisruptsell membrane components. Researchers found there are cannabinoid receptors located in the brain and body cells. The specific action of this cannabinoid suggests that delta 9-THC inhibits adenylate cyclase activity in N18TG2 neuroblastoma cells cultured in vitro and the use of radiolabeled analogs alloanalogstection of brain-specific cannabinoid sites. There are two types of cannabinoid receptors (CB) namely CB1 in the central nervous system and CB2 in immune system cells. Stimulation of these cannabinoid receptors can be found in the frontal cortex, basal ganglia, cerebellum, and hippocamp, u This stimulation causes the release of neurotransmitters that have a relaxing effect, euphoria, increased self-confidence, cardiovascular complications, peripheral disorders,d neurology,ical complications.

##### d) Poor Diet

Unfavorable lifestyles such as consuming foods high in fat and cholesterol, lack of movement or physical activity, and exercise can trigger an increase in stroke at a young age (Budi et al., 2020). Foods that are not good and have a high risk of stroke include meat, egg yolk,s, and excessive salt intake. Eating foods that contain B vitamins and following a Mediterranean diet (consuming fruits, vegetables, nuts, and seeds) can reduce risk factors for stroke at a young age (Spence, 2019).

##### e) Low Activity

The factor of lack of exercise, people who are sedentary can lead to increased blood pressure, weight gain, and atherosclerosis (Budi et al., 2020). In addition, low physical activity and a sedentary lifestyle are risk factors for stroke at a young age. A study stated that low physical activity (walking less than 1 mile/day) was a risk factor for stroke at a young age for both men and women with the percentages of 46.6% and 50.4%, respectively. However, work or physical activity that requires standing for too long also increases the risk of stroke. So it is necessary to have a balanced physical activity as recommended by the American Heart Association by exercising 2.5 hours per week to significantly reduce the risk of stroke (Polivka et al., 2019).

##### f) Use of Oral Contraceptives in Women

The use of contraceptive drugs has a very high risk of ischemic stroke in young women, especially in women who have migraines and use contraceptive drugs (CHC). The use of this dose of CHC resulted in a decrease in estrogen levels which resulted in ischemic stroke in young women (Ornello et al., 2020). Likewise, according to Polivka et al (2019), the use of oral contraceptives is associated with an increased risk of ischemic stroke in young women, especially the use of estrogen-based contraceptives in the form of piano containing > 50 grams of estrogen. The mechanism of SHC increases the risk of stroke due to its prothrombotic and proinflammatory effects. Another study states that estrogen is very influential on the rate of stroke in humans. Progesterone according to some evidence can also be neuroprotective after traumatic brain injury.

The etiology of stroke in contraceptives results from a lower intracranial and extracranial atheromatous and a higher incidence of vocal hypoperfusion in CTP. The mechanism of stroke in this systemic hormonal contraceptive is the prothrombotic state of the arteries in paradoxical embolization or accelerated atherosclerosis (Correia et al., 2021).

#### d. Age factor

Stroke at a young age is closely related to the age factor, with increasing age, the risk of stroke increases exponentially. A study explains that there is an increased incidence of ischemic stroke at the age of < 35 years and < 30 years (Ekker et al., 2019). This happens because the older you get, the system declines in the blood vessels (Budi & Bahar, 2017). The research of Wu et al (2021) based on the random survival forest analysis showed that age is a strong predictor of ischemic stroke.

#### e. Gender Factor

The results of the study stated that gender factor greatly influences the incidence of stroke at a young age. The male gender has a higher risk factor than the female. Research study data states that 88.3% of male patients have a risk factor for ischemic stroke at a higher age than women. The prevalence of comorbidities was significantly higher in males (M. Tang et al., 2020).

Researcher Dr.Sharadha et al (2020) stated the same thing that men have a higher risk of stroke at a young age due to a man’s poor lifestyle such as consuming alcohol and smoking. In addition, the role and duties of a man are very important, especially in terms of family and society which increase stress and depression (Alchuriyah & Wahjuni, 2016)

## 4. CONCLUSION

Stroke today does not only attack at old age but also at a young ageagehndiratta & Mehndiratta (2021) stated that stroke at a young age is one of the causes of morbidity and mortality that has an impact on disability and reduced physical productivity at a young age. Young strostrokengup was mostly found at the age of 15-45 years or <55 years. The increase in stroke at a young age is due to several factors, one of which is the risk factor for stroke at a young age. So the purpose of this study is to detail several risk factors for stroke at a young age into general factors.

The factors, in general, include 5 factors that influence the incidence of stroke at a young age, namely physical factors, psychological factors, lifestyle factors, age factors, and gender factors. Of these factors, two factohavevee the highest percentage, namely physical factors and lifestyle factors with percentages of 85% and 55%, respectively. The age factor is the lowest in the study of stroke incidence at a young age.

## Data Availability

All data produced in the present work are contained in the manuscript

## IMPLICATION OF RESULTS ON NURSING PRACTICE

This literature review study is expected to be used as input for nursing science, especially medical-surgical nursing, in opening new insights about the trend of a stroke at a young age to be at the forefront of preventive and promotive efforts to improve health and well-being.

## REFERENCES

Aguilera-Pena, M. P., Cardenas-Cruz, A. F., Baracaldo, I., Garcia-Cifuentes, E., Ocampo-Navia, M. I., & Coral, E. J. (2021). Ischemic stroke in young adults in Bogota, Colombia: a cross-sectional study. Neurological Sciences, 42(2), 639–645. https://doi.org/10.1007/s10072-020-04584-2

Aigner, A., Grittner, U., Rolfs, A., Norrving, B., Siegerink, B., & Busch, M. A. (2017). Contribution of Established Stroke Risk Factors to the Burden of Stroke in Young Adults. Stroke, 48(7), 1744–1751. https://doi.org/10.1161/STROKEAHA.117.016599

Akinwuntan, A., Hu, X., Terrill, A. L., Burns, S. P., Hay, C. C., & Belagaje, S. R. (2021). Young Stroke: Resources for Patients, Their Families, and Caregivers for Long-Term Community Living. Archives of Physical Medicine and Rehabilitation, 102(5), 1035–1039. https://doi.org/10.1016/j.apmr.2020.10.108

Alchuriyah, S., & Wahjuni, C. U. (2016). Faktor Risiko Kejadian Stroke Usia Muda Pada Pasien Rumah Sakit Brawijaya Surabaya. Jurnal Berkala Epidemiologi, 4(1), 62–73. https://doi.org/10.20473/jbe.v4i1.62-73

Alloubani, A., Saleh, A., & Abdelhafiz, I. (2018). Hypertension and diabetes mellitus as a predictive risk factor for stroke. Diabetes and Metabolic Syndrome: Clinical Research and Reviews, 12(4), 577–584. https://doi.org/10.1016/j.dsx.2018.03.009

Anggriani, A., Zulkarnain, Z., Sulaiman, S., & Gunawan, R. (2018). Pengaruh Rom (Range Of Motion) Terhadap Kekuatan Otot Ekstremitas Pada Pasien Stroke Non Hemoragic. Jurnal Riset Hesti Medan Akper Kesdam I/BB Medan, 3(2), 64. https://doi.org/10.34008/jurhesti.v3i2.46

Bartholomé, L., & Winter, Y. (2020). Quality of Life and Resilience of Patients With Juvenile Stroke: A Systematic Review. Journal of Stroke and Cerebrovascular Diseases, 29(10). https://doi.org/10.1016/j.jstrokecerebrovasdis.2020.105129

Budi, H., & Bahar, I. (2017). Faktor Resiko Stroke Hemorragic Pada Pasien Usia Produktif. Jurnal Sehat Mandiri, 12(2), 29–36.

Budi, H., Bahar, I., & Sasmita, H. (2020). Faktor Risiko Stroke Pada Usia Produktif Di Rumah Sakit Stroke Nasional (Rssn) Bukit Tinggi. Jurnal Persatuan Perawat Nasional Indonesia (JPPNI), 3(3), 129. https://doi.org/10.32419/jppni.v3i3.163

Cho, S., & Yang, J. (2018). What do experimental models teach us about comorbidities in stroke? Stroke, 49(2), 501–507. https://doi.org/10.1161/STROKEAHA.117.017793

Cipolla, M. J., Liebeskind, D. S., & Chan, S. L. (2018). The importance of comorbidities in ischemic stroke: Impact of hypertension on the cerebral circulation. Journal of Cerebral Blood Flow and Metabolism, 38(12), 2129–2149. https://doi.org/10.1177/0271678X18800589

Corraini, P., Szépligeti, S. K., Henderson, V. W., Ording, A. G., Horváth-Puhó, E., & Sørensen, H. T. (2018). Comorbidity and the increased mortality after hospitalization for stroke: a population-based cohort study. Journal of Thrombosis and Haemostasis, 16(2), 242–252. https://doi.org/10.1111/jth.13908

Correia, P., Machado, S., Meyer, I., Amiguet, M., Eskandari, A., & Michel, P. (2021). Ischemic stroke on hormonal contraceptives: Characteristics, mechanisms, and outcome. European Stroke Journal, 6(2), 205–212. https://doi.org/10.1177/23969873211019586

Coupland, A. P., Thapar, A., Qureshi, M. I., Jenkins, H., & Davies, A. H. (2017). The definition of stroke. Journal of the Royal Society of Medicine, 110(1), 9–12. https://doi.org/10.1177/0141076816680121

De Giuli, V., Grassi, M., Lodigiani, C., Patella, R., Zedde, M., Gandolfo, C., Zini, A., DeLodovici, M. L., Paciaroni, M., Del Sette, M., Azzini, C., Toriello, A., Musolino, R., Calabrò, R. S., Bovi, P., Sessa, M., Adami, A., Silvestrelli, G., Cavallini, A., … Pezzini, A. (2017). Association Between Migraine and Cervical Artery Dissection The Italian Project on Stroke in Young Adults. JAMA Neurology, 74(5), 512–518. https://doi.org/10.1001/jamaneurol.2016.5704

Dguzeh, U., Haddad, N. C., Smith, K. T. S., Johnson, J. O., Doye, A. A., Gwathmey, J. K., & Haddad, G. E. (2018). Alcoholism: A multi-systemic cellular insult to organs. International Journal of Environmental Research and Public Health, 15(6). https://doi.org/10.3390/ijerph15061083

Dinh, P. C., Schrader, L. A., Svensson, C. J., Margolis, K. L., Silver, B., & Luo, J. (2019). Smoking cessation, weight gain, and risk of stroke among postmenopausal women. Preventive Medicine, 118(October 2018), 184–190. https://doi.org/10.1016/j.ypmed.2018.10.018

Dr.Sharadha, S., B.Nadia, S., C. Vamsi, M., K.Sandeep, R., & S., K. (2020). Risk Factors Associated With Cerebro-Vascular Accident Ischemic Stroke In Young And Elderly Population. International Journal of Pharma and Bio Sciences, 10(4). https://doi.org/10.22376/ijpbs/lpr.2020.10.4.p13-21

Dyussenbayev, A. (2017). Age Periods Of Human Life. Advances in Social Sciences Research Journal, 4(6), 258–263. https://doi.org/10.14738/assrj.46.2924

Echouffo-Tcheugui, J. B., Xu, H., Matsouaka, R. A., Xian, Y., Schwamm, L. H., Smith, E. E., Bhatt, D. L., Hernandez, A. F., Heidenreich, P. A., & Fonarow, G. C. (2018). Diabetes and long-term outcomes of ischaemic stroke: Findings from get with the guidelines-stroke. European Heart Journal, 39(25), 2376–2386. https://doi.org/10.1093/eurheartj/ehy036

Ekker, M. S., Boot, E. M., Singhal, A. B., Tan, K. S., Debette, S., Tuladhar, A. M., & de Leeuw, F. E. (2018). Epidemiology, etiology, and management of ischaemic stroke in young adults. The Lancet Neurology, 17(9), 790–801. https://doi.org/10.1016/S1474-4422(18)30233-3

Ekker, M. S., Verhoeven, J. I., Vaartjes, I., Van Nieuwenhuizen, K. M., Klijn, C. J. M., & De Leeuw, F. E. (2019). Stroke incidence in young adults according to age, subtype, sex, and time trends. Neurology, 92(21), e2444–e2454. https://doi.org/10.1212/WNL.0000000000007533

Gaffey, A. E., Rosman, L., Burg, M. M., Haskell, S. G., Brandt, C. A., Skanderson, M., Dziura, J., & Sico, J. J. (2021). Posttraumatic Stress Disorder, Antidepressant Use, and Hemorrhagic Stroke in Young Men and Women: A 13-Year Cohort Study. Stroke, January, 121–129. https://doi.org/10.1161/STROKEAHA.120.030379

Hathidara, M. Y., Saini, V., & Malik, A. M. (2019). Stroke in the Young: a Global Update. Current Neurology and Neuroscience Reports, 19(11), 1–8. https://doi.org/10.1007/s11910-019-1004-1

Karunia., E. (2016). Hubungan antara dukungan keluarga dengan kemandirian Activity of Daily Living Pascastroke. July, 213–224. https://doi.org/10.20473/jbe.v4i2.2016.213

Marbun, J. T. Seniman, & Andayani, U. (2018). Classification of stroke disease using convolutional neural network. Journal of Physics: Conference Series, 978(1). https://doi.org/10.1088/1742-6596/978/1/012092

Matuja, S. S., Munseri, P., & Khanbhai, K. (2020). The burden and outcomes of stroke in young adults at a tertiary hospital in Tanzania: A comparison with older adults. 1–10. https://doi.org/10.21203/rs.3.rs-16407/v2

Mehndiratta, M., & Mehndiratta, P. (2021). Stroke in the young: Newer concepts in etiopathogenesis and risk factors. Astrocyte, 5(1), 26–32. https://doi.org/10.4103/astrocyte.astrocyte_48_18

Melangi, S. (2020). Klasifikasi Usia Berdasarkan Citra Wajah Menggunakan Algoritma Artificial Neural Network dan Gabor Filter. Jambura Journal of Electrical and Electronics Engineering, 2(2), 60–67. https://doi.org/10.37905/jjeee.v2i2.6956

Millah, F. N., Uyun, Q., & Sulistyarini, I. R. (2020). Pelatihan shalat khusyuk meningkatkan kebahagiaan pada family caregiver pasien stroke. Jurnal Intervensi Psikologi, 12(2), 81–94.

Nelson, M. L. A., McKellar, K. A., Yi, J., Kelloway, L., Munce, S., Cott, C., Hall, R., Fortin, M., Teasell, R., & Lyons, R. (2017). Stroke rehabilitation evidence and comorbidity: A systematic scoping review of randomized controlled trials. Topics in Stroke Rehabilitation, 24(5), 374–380. https://doi.org/10.1080/10749357.2017.1282412

Nutakki, A., Chomba, M., Chishimba, L., Zimba, S., Gottesman, R. F., Bahouth, M. N., & Saylor, D. (2021). Risk factors and outcomes of hospitalized stroke patients in Lusaka, Zambia. Journal of the Neurological Sciences, 424(March), 117404. https://doi.org/10.1016/j.jns.2021.117404

Oktari, I., Febtrina, R., Malfasari, E., & Guna, S. D. (2020). Tingkat Ketergantungan Dalam Pemenuhan Aktivitas Sehari Hari Berhubungan dengan Harga Diri Penderita Stroke. Jurnal Ilmiah Permas: Jurnal Ilmiah STIKES Kendal, 10(2), 185–194.

Ornello, R., Canonico, M., Merki-Feld, G. S., Kurth, T., Lidegaard, Ø., MacGregor, E. A., Lampl, C., Nappi, R. E., Martelletti, P., & Sacco, S. (2020). Migraine, low-dose combined hormonal contraceptives, and ischemic stroke in young women: a systematic review and suggestions for future research. Expert Review of Neurotherapeutics, 20(4), 313–317. https://doi.org/10.1080/14737175.2020.1730816

Owolabi, M. O., Sarfo, F., Akinyemi, R., Gebregziabher, M., Akpa, O., Akpalu, A., Wahab, K., Obiako, R., Ovbiagele, B., Sarfo, F. S., Akinyemi, R., Gebregziabher, M., Akpa, O., Akpalu, A., Obiako, R., Ovbiagele, B., Tiwari, H. K., Arnett, D., Lackland, D., … Owolabi, L. (2018). Dominant modifiable risk factors for stroke in Ghana and Nigeria (SIREN): a case-control study. The Lancet Global Health, 6(4), e436–e446. https://doi.org/10.1016/S2214-109X(18)30002-0

Parekh, T., Pemmasani, S., & Desai, R. (2020a). Marijuana Use Among Young Adults (18–44 Years of Age) and Risk of Stroke A Behavioral Risk Factor Surveillance System Survey Analysis. Stroke, 51(1), 308–310. https://doi.org/10.1161/STROKEAHA.119.027828

Parekh, T., Pemmasani, S., & Desai, R. (2020b). Risk of Stroke With E-Cigarette and Combustible Cigarette Use in Young Adults. American Journal of Preventive Medicine, 58(3), 446–452. https://doi.org/10.1016/j.amepre.2019.10.008

Pedersen, M. G. B., Olsen, M. S., Schmidt, M., Johnsen, S. P., Learn, C., Laursen, H. B., & Madsen, N. L. (2019). Ischemic Stroke in Adults With Congenital Heart Disease: A Population-Based Cohort Study. Journal of the American Heart Association, 8(15). https://doi.org/10.1161/JAHA.118.011870

Polivka, J., Polivka, J., Pesta, M., Rohan, V., Celedova, L., Mahajani, S., Topolcan, O., & Golubnitschaja, O. (2019). Risks associated with the stroke predisposition at a young age: facts and hypotheses in light of individualized predictive and preventive approach. EPMA Journal, 10(1), 81–99. https://doi.org/10.1007/s13167-019-00162-5

Prasad, M., Khanna, P., Katyal, V. K., & Verma, R. (2020). Acute Psychological Stress is a Trigger for Stroke: A Case-Crossover Study. Journal of Stroke and Cerebrovascular Diseases, 29(6), 104799. https://doi.org/10.1016/j.jstrokecerebrovasdis.2020.104799

Putri, V. A., & Muti, A. F. (2017). Profil Penggunaan Neuroprotektor pada Pasien Stroke Iskemik di RSPAD Gatot Soebroto Jakarta. Sainstech Farma, 10(1), 13–20.

Rashid, M. H. U., Kashem, M. A., Biswas, S., & Hoque, M. M. (2020). Risk factors in young stroke. Journal of Medicine (Bangladesh), 21(1), 26–30. https://doi.org/10.3329/jom.v21i1.44097

Rosman, L., Sico, J. J., Lampert, R., Gaffey, A. E., Ramsey, C. M., Dziura, J., Chui, P. W., Cavanagh, C. E., Brandt, C., Haskell, S., & Burg, M. M. (2019). Posttraumatic Stress Disorder and Risk for Stroke in Young and Middle-Aged Adults A 13-Year Cohort Study. 1–8. https://doi.org/10.1161/STROKEAHA.119.026854

Sarecka-Hujar, B., & Kopyta, I. (2020). Risk factors for recurrent arterial ischemic stroke in children and young adults. Brain Sciences, 10(1), 1–20. https://doi.org/10.3390/brainsci10010024

Setiawan, Y. (2018). Faktor – Faktor yang Berhubungan dengan Kejadian Stroke Pada Usia Muda di Ruang Wijaya RSUD Kota Bekasi. Jurnal Ilmiah Keperawatan, 1–12. http://jurnal.imds.ac.id/imds/index.php/JIKep/article/download/71/68

Shihmanter, R., Schattner, A., Friedman, J., Kushner, N., & Miller, E. B. (2021). Prospective observational study of young adult ischemic stroke patients. April, 1–9. https://doi.org/10.1002/brb3.2283

Si, Y., Xiang, S., Zhang, Y., Lu, T., Guo, J., Xiao, X., & Dong, L. (2020). Clinical pro fi le of aetiological and risk factors of young adults with ischemic stroke in West China. Clinical Neurology and Neurosurgery, 193(February), 105753. https://doi.org/10.1016/j.clineuro.2020.105753

Sinaga, J., & Sembiring, E. (2019). Pencegahan Stroke Berulang Melalui Pemberdayaan Keluarga Dan Modifikasi Gaya Hidup. Jurnal Abdimas, 22(2), 143–150.

Spence, J. D. (2019). Nutrition and risk of stroke. Nutrients, 11(3). https://doi.org/10.3390/nu11030647

Stack, C. A., & Cole, J. W. (2018). Ischemic stroke in young adults. Current Opinion in Cardiology, 33(6), 594–604. https://doi.org/10.1097/HCO.0000000000000564

Stein, J. (2011). Stroke. In Essentials of Physical Medicine and Rehabilitation (Fourth Edi). Elsevier Inc. https://doi.org/10.1016/B978-0-323-54947-9.00159-0

Stephen, F., Opare-sem, O., Agyei, M., Akassi, J., Owusu, D., Owolabi, M., & Ovbiagele, B. (2018). Risk factors for stroke occurrence in a low HIV endemic West African country : A case-control study. Journal of the Neurological Sciences, 395(September), 8–16. https://doi.org/10.1016/j.jns.2018.09.021

Susilawati, F., & Nurhayati, H. K. (2018). Faktor Resiko Kejadian Stroke di Rumah Sakit. Jurnal Keperawatan, 14(1), 41–48.

Syifa, N., Amalia, L., Yulianti Bisri, D., Kedokteran Universitas Padjadjaran, F., Neurologi Fakultas Kedokteran Universitas Padjadjaran, D., Sakit Hasan Sadikin, R., Anestesiologi dan Terapi Intensif, D., & Kedokteran Universitas Padjadjaran-Rumah Sakit Hasan Sadikin, F. (2017). Gambaran Epidemiologi Pasien Stroke Dewasa Muda yang Dirawat di Bangsal Neurologi RSUP Dr.Hasan Sadikin Bandung Periode 2011–2016. Jurnal Neuroanestesi Indonesia, 6(3), 143–150. http://www.inasnacc.org/images/vol6no03Oktober2017/Volume06Nomor03Oktober2017NadyaSyifa.pdf

Tang, E. Y. H., Price, C., Stephan, B. C. M., Robinson, L., & Exley, C. (2020). Impact of Memory Problems Post-stroke on Patients and Their Family Carers: A Qualitative Study. Frontiers in Medicine, 7(June). https://doi.org/10.3389/fmed.2020.00267

Tang, M., Yao, M., Zhu, Y., Peng, B., Zhou, L., & Ni, J. (2020). Sex differences of ischemic stroke in young adults—A single-center Chinese cohort study. Journal of Stroke and Cerebrovascular Diseases, 29(9), 1–6. https://doi.org/10.1016/j.jstrokecerebrovasdis.2020.105087

Tun, N. N., Arunagirinathan, G., Munshi, S. K., & Pappachan, J. M. (2017). Diabetes mellitus and stroke: A clinical update. World Journal of Diabetes, 8(6), 235. https://doi.org/10.4239/wjd.v8.i6.235

van Alebeek, M. E., Arntz, R. M., Ekker, M. S., Synhaeve, N. E., Maaijwee, N. A. M. M., Schoonderwaldt, H., van der Vlugt, M. J., van Dijk, E. J., Rutten-Jacobs, L. C. A., & de Leeuw, F. E. (2018). Risk factors and mechanisms of stroke in young adults: The FUTURE study. Journal of Cerebral Blood Flow and Metabolism, 38(9), 1631–1641. https://doi.org/10.1177/0271678X17707138

Villa, R. F., Ferrari, F., & Moretti, A. (2018). Post-stroke depression: Mechanisms and pharmacological treatment. Pharmacology and Therapeutics, 184(XXXX), 131–144. https://doi.org/10.1016/j.pharmthera.2017.11.005

Wu, Z., Huang, Z., Lichtenstein, A. H., Liu, Y., Chen, S., Jin, Y., Na, M., Bao, L., Wu, S., & Gao, X. (2021). The risk of ischemic stroke and hemorrhagic stroke in Chinese adults with low-density lipoprotein cholesterol concentrations < 70 mg/dL. BMC Medicine, 19(1), 1–12. https://doi.org/10.1186/s12916-021-02014-4

Yahya, T., Jilani, M. H., Khan, S. U., Mszar, R., Hassan, S. Z., Blaha, M. J., Blankstein, R., Virani, S. S., Johansen, M. C., Vahidy, F., Cainzos-Achirica, M., & Nasir, K. (2020). Stroke in young adults: Current trends, opportunities for prevention and pathways forward. American Journal of Preventive Cardiology, 3(June), 100085. https://doi.org/10.1016/j.ajpc.2020.100085

Yasin Abanoz, Yeşim Gülen Abanoz, Ayşegü, Gündüz, Derya Uludüz, Birsen İnce, Burcu Yavuz, B. G. (2017). Migraine as a risk factor for young patients with ischemic stroke : a case-control study. Neurol Sci. https://doi.org/10.1007/s10072-017-2810-3

Yaslina, Y., Maidaliza, M., & Hayati, I. (2019). Pengaruh Pemberian Discharge Planning Terhadap Kemampuan Keluarga Dalam Perawatan Pasca Stroke Di Rumah Tahun 2019. JURNAL KESEHATAN PERINTIS (Perintis’s Health Journal), 6(1), 54–59. https://doi.org/10.33653/jkp.v6i1.240

Zhang, N., Zhang, L., Wang, Q., Zhao, J., Liu, J., & Wang, G. (2020). Cerebrovascular risk factors associated with ischemic stroke in a young non-diabetic and non-hypertensive population : a retrospective case-control study. 1–8.

Zhang, Q., Liu, Y., Jiang, M., Liu, Y., Gu, S., Tong, H., & Liu, H. (2020). Temporal Trends in the Risk Factors and Clinical Characteristics of Ischemic Stroke in Young Adults. Journal of Stroke and Cerebrovascular Diseases, 29(8), 1–7. https://doi.org/10.1016/j.jstrokecerebrovasdis.2020.104914

